# Work Expectations, Employment Transitions, and Psychological Distress During the Great Recession Among Older Americans and South Koreans

**DOI:** 10.1101/2024.11.21.24317724

**Authors:** Linh Dang, Toni Antonucci, Carlos Mendes de Leon, Briana Mezuk

**Affiliations:** Center for Social Epidemiology and Population Health, Department of Epidemiology, University of Michigan School of Public Health, Ann Arbor, Michigan, USA; Department of Psychology, University of Michigan, Ann Arbor, MI USA; Department of Global Health, Georgetown University School of Health, Washington, DC, USA; Institute for Social Research, University of Michigan, Ann Arbor, MI USA

**Keywords:** retirement, economic crisis, cross-country comparison, longitudinal

## Abstract

**Background and objectives:** Perceived uncertainty about future employment and work transitions during economic crises can negatively affect mental health of workers. This study examined the association between work expectations and psychological distress among “Baby Boomers” (born 1948-1965) in the United States and South Korea and explored how employment transitions moderated this association before, during, and after the Great Recession.

**Research Design and Methods:** Data came from 2006-2018 waves of the Health and Retirement Study (n=2,647) and Korean Longitudinal Study of Aging (n=1,454). Perceived expectations of working in the next five years were reported on a probability scale (0-100%). Psychological distress was assessed by the Center for Epidemiologic Studies Depression Scale. Six mutually-exclusive employment transitions during the Recession were constructed: entrance to full-time employment, precarious employment, stable full-time employment, self-employment, exit from the labor force, and consistently out of the labor force. Multivariate mixed-effects logistic regression models were used to assess the association between work expectations, employment transitions, and psychological distress.

**Results:** Higher work expectations were associated with lower odds of psychological distress; this association was most robustly observed during the recession (OR_US_=0.93 [0.89, 0.98], OR_Korea_=0.92 [0.84, 1.01]) and recovery (OR_US_=0.95 [0.92, 0.98], OR_Korea_=0.84 [0.80, 0.89]). The relationship between expectations and distress was pronounced among respondents who were self-employed or transitioning into self-employment. Findings were broadly similar comparing two countries.

**Discussion and Implications:** Expectations regarding, and transitions in, employment impact psychological distress. Findings emphasize the need to support the mental health of older adults nearing retirement during periods of economic crisis.

## INTRODUCTION

The Great Recession (“Recession” thereafter), an 18-month period of substantial global economic contraction, witnessed unprecedented rates of layoffs and unemployment, with more than 205 million unemployed workers in 2009 (an increase of 27 million since 2007) in the United States (US) alone (United Nations, 2011). The Recession had significant impacts on the “Baby Boom” generation (born 1948 – 1965) as this cohort experienced the crisis during the period of their working lives directly preceding typical retirement (ages 60-65).

Compared with other cohorts, Baby Boomers experienced the largest decline in retirement income and wealth during the Recession (Gustman et al., 2012), partly attributable to losses in their defined contribution (DC) retirement plans (e.g., 401k) (Munnell & Rutledge, 2013). Unlike pensions which provide a fixed, pre-determined retirement benefits, DC plans depend on the deposits made by employees into individual retirement accounts. However, workers often stop or reduce contributions to DC plans in response to changes in employment, including during the Recession (Dushi et al., 2013). Falling equity values and interest rates also impacted balances of DC plans; indeed, in 2010, the combined balance of household 401ks and individual saving accounts were only $120,000 for Americans aged 55-65 (Munnell & Rutledge, 2013). The financial consequences of the Recession on DC plans uniquely impacted Baby Boomers because they were the first cohort to broadly depend on 401k and similar DC retirement plans rather than defined-benefit plans (US Bureau of Labor Statistics, 2021).

### Impacts of the Recession on employment decisions and patterns among Baby Boomers

The Recession negatively impacted the retirement resources of Baby Boomers in a manner that influenced their retirement decisions. Many of the oldest in this cohort postponed retirement and remained in the labor force (Population Reference Bureau, 2015), presumably to counter losses in retirement wealth (Kalleberg & Von Wachter, 2017). Indeed, despite rising unemployment rates, the labor force participation for adults aged 62-64 increased between 2007 and 2009 (Johnson & Butrica, 2012). Beyond material job transitions, the Recession also impacted the expectations regarding future employment; the number of US adults expecting to retire before age 65 declined from 33% to 23% between 2006 and 2011 (Helman et al., 2011). This expectancy of delaying retirement was strongly correlated with prior experience of wealth shocks (Coile & Levine, 2015), losses in retirement savings (Szinovacz et al., 2014), and pessimism about the future stock market returns (McFall, 2011).

Despite the intention to work longer, approximately 15% of older workers were unemployed during the Recession, with tripled unemployment rate among those over age 62 (Johnson & Butrica, 2012). Further, older adults who lost their jobs faced greater difficulty in regaining employment than younger workers. Half of unemployed US adults aged 50-61 took more than nine months to find another job, as compared to only six months among adults aged 25-34, during and after the Recession (Johnson & Butrica, 2012). Many eventually accepted part-time jobs (Kalleberg & Von Wachter, 2017), self-employment (Beckhusen, 2014b), or lower-paying, temporary work (Benach et al., 2014) due to the difficulty in finding full-time employment. In 2010, the share of part-time employed adults was double compared to pre-recession, with approximately 10% of Americans over age 50 being under- or part-time employed (Johnson & Butrica, 2012).

### Conceptual frameworks linking economic recessions to mental health

A well-established body of research has documented the negative impacts of economic recessions on the mental health of older adults (Frasquilho et al., 2016). Many of these studies reported significant increases in psychological distress among older adults during and after recessions (S. Lee et al., 2010; Pruchno et al., 2017). Recessions amplify several risk factors for psychological distress, such as unemployment, financial strains, and wealth losses (Kalleberg & Von Wachter, 2017). Importantly, perceived economic insecurity heightened by recessions may have even more detrimental consequences on mental health than actual job loss. Workers who perceived job insecurity had significantly worse emotional distress, even after accounting for objective employment security (Burgard et al., 2009). This suggests that beyond the actual employment status, concerns about economic uncertainty, particularly those regarding future employment, play an important role in influencing the psychological well-being of older adults during recessionary periods.

In line with this research on job insecurity and mental health, *expectations* about future employment are also associated with mental health among older adults (Abrams et al., 2021; Mezuk et al., 2022). These expectations are subjective assessments about the likelihood of future events, typically assessed on a probability scale (e.g., 0% to 100% chance something will happen) and are related to several other future-oriented psychosocial constructs, including hopelessness and anxiety (Liu et al., 2015). Both hopelessness and anxiety are established risk factors for psychological distress via connections with perceived uncertainty and negative thoughts about the future in the context of the economic recessions (Parada-Fernández et al., 2020). Specifically, the construct of *uncertainty* relects (in)flexibility or (un)willingness to adapt to new situations and/or recognition that factors outside one’s control can impact likelihood of future events (Hershfield, 2011). Indeed, cognitive resources such as *perceived control* over stressful life events appear to moderate the risk for economic-related psychological distress (Frazier et al., 2011).

### Exploring variations in the mental health correlates of the Recession

Although the Recession increased the prevalence of psychological distress overall (Frasquilho et al., 2016), its mental health impacts were heterogeneous. Some individuals may experience increases in distress symptoms, while others remain unchanged or even experience declines in symptoms (Pruchno et al., 2017). This variability is partially explained by the *cumulative inequality theory*, a framework that integrates the life course perspective, cumulative (dis)advantage, and stress process theories (Ferraro & Shippee, 2009). Based on this theory, differential exposure to recession-related stressors and psychological vulnerability may arise from an accumulation of socioeconomic disadvantages (“stressors”) in earlier life, which are subsequently exacerbated by the Recession and then adversely influence mental health (Crystal et al., 2017). Moreover, principles from the cumulative (dis)advantage emphasizes that experiences of stress may span multiple life domains (e.g., employment, finance, and housing) and may diffuse or interact with those from other domains (Burgard et al., 2013). For example, involuntary job loss not only heightens the risks for psychological distress directly (Riumallo-Herl et al., 2014) but also leads to a range of secondary stressors such as declines in household income and wealth, losses in health insurance coverage and pensions, and housing instability (Burgard et al., 2013).

One source of inequality exacerbated by the Recession entailed differences in exposure to economic adversity (Koltai & Stuckler, 2019). While unemployment and unexpected job loss are frequently documented in the recession literature, there is a considerable heterogeneity in these experience across sectors and individual workers. Previous studies have identified several common employment transitions during the Recession, including stable employment, precarious or temporary employment, consistently being out of the labor force, transitioning out of the labor force, and intermittent employment (Kalousová & Burgard, 2022; J. Lee & Kim, 2017). Moreover, individuals in different employment transitions experience different stressors and subsequently differ in liability for psychological distress (Thomas et al., 2005). In particular, transitions to precarious/temporary employment were associated with heightened feelings of job insecurity and elevated risk of distress (Kalousová & Burgard, 2022). Overall, these findings emphasize the need to better understand how employment transitions may moderate the effect of the Recession on mental health among older workers.

### Cross-country differences in the impacts of the Recession

While the Recession disrupted the economy and financial market worldwide, the scale and timing vary across countries, with the most severe impacts in the US (Arias & Wen, 2015). At the height of the Recession, the US unemployment rate was nearly doubled, from 5% in December 2007 to 10% in October 2009. In contrast, financial reforms and policies, especially those implemented following the Asian financial crisis of the late 1990s, helped buffer the impact of the Recession in many Asian economies (Bernanke, 2009). Indeed, many Asian countries experienced minimal impact from the Recession, retaining relatively low unemployment rates and substantial GDP growth throughout the crisis (Arias & Wen, 2015). Moreover, even those Asian economies that were affected recovered from the Recession rather quickly, showing strong economic growth by the second quarter of 2010 (Arias & Wen, 2015); in contrast, US and European countries took nearly eight years to return to pre-recession growth (Weinberg, 2013). Therefore, cross-country comparisons can provide insights into how the sociocultural, political, and macro-economic contexts in which recessions occur may shape the association between work expectations and mental health.

### Present study

While previous research showed that subjective expectations regarding future employment were associated with mental health among older adults, it is unclear whether employment transitions, particularly during a period of crisis like the Recession, may moderate this association. To that end, this study explored two research questions:

1. How does the association between work expectations and psychological distress vary across three time periods of the Recession: pre-recession (2006-2007), recession (2008-2009), and recovery (2010-2018)?
2. How do employment transitions moderate the association between work expectations and psychological distress during the three time periods of the Recession?

This study focused on the “Baby Boom” generation given the unique impact of the Recession on retirement resources of this cohort. Moreover, the associations between work expectations, employment transitions, and psychological distress were explored in the US and South Korea (“Korea” thereafter), two countries that differed substantially in the scale of the Recession. We hypothesized that the associations between work expectations and psychological distress will be stronger during the Recession than other periods, and this association will be moderated by employment transitions. Further, the effects of the Recession and employment transitions will differ between the US and Korea.

## METHODS

### Data source and study population

Data came from two population-based longitudinal studies of aging conducted in the US and Korea, including the Health and Retirement Study (HRS, n∼20,000 respondents ages 51+ surveyed biennially since 1992) and the Korean Longitudinal Study of Aging (KLoSA, n∼10,000 respondents ages 45+ surveyed biennially since 2006). Both the HRS and KLoSA employ a complex, multi-stage study design that enrolls refreshment cohorts (every six years in the HRS and at the 2014 wave in KLoSA) to remain representative of the contemporaneous older adult population. These harmonized studies provide a unique opportunity to investigate how expectations for late-life employment relate to psychological distress within the economic context of the Recession in the US and Korea. Additional details on the HRS and KLoSA are described elsewhere (Joon et al., 2007; Sonnega et al., 2014).

This study focused on two cohorts: Early Baby Boomers in the HRS (born 1948 – 1953) and Baby Boomers in the original KLoSA sample (born 1955 – 1963). Other Baby Boom cohorts in the HRS (e.g., Mid and Late Baby Boomers) and the refreshment sample of KLoSA were excluded because these respondents were not interviewed during the Recession. The study period spanned 12 years (waves 8-14 of HRS and waves 1-7 of KLoSA), with the 2006 wave serving as the baseline and the 2018 wave as the last follow-up. The analytical sample included 4,101 (2,647 in the HRS and 1,454 in KLoSA) non-proxy respondents who had available data on labor force status between interview waves 2006 and 2010 as well as complete data on work expectations, psychological distress, and all potential covariates for at least one wave from 2006 to 2018. These interview waves were chosen because 2006 was the first interview wave of KLoSA, and 2018 was the latest available interview wave prior to the COVID-19 pandemic. While the pandemic may affect the association between work expectations and psychological distress, its impact is outside the scope of the present study.

Supplementary Figure 1 details the analytic sample selection process. Differences between the analytic sample and excluded respondents were summarized in Supplementary Table 1; most relevant to this investigation is that excluded respondents were older, more likely to have lower socioeconomic status, and in poorer health compared to the analytic sample.

The HRS and KLoSA are approved by the research ethics review committees in their respective countries, and all respondents are provided written informed consent. This analysis used only publicly available data (https://g2aging.org) and was exempt from human subject regulation.

### Work expectations

Questions on work expectations were asked at every interview wave in both the HRS and KLoSA; however, there are several key differences between the two datasets. First, the KLoSA assessed work expectations by a single question asking the respondents about their subjective likelihood of working “at the present job in the next 5 years.” Specifically, respondents younger than 50 were asked about the likelihood of working after age 55; respondents aged 50-54 were asked about working after age 60, respondents aged 55-59 were asked about working after age 65; and finally, respondents aged 60+ were asked about the likelihood of working in the next 5 years. On the other hand, the HRS assessed work expectations by three questions asking the respondents about the subjective likelihood of “working full-time” after age 62, 65, and 70.

These questions were then used to create a new variable based on the respondents’ age: expectations of working after age 62 were used for respondents aged 56 or younger; expectations of working after age 65 for those aged 57-62; and expectations of working after age 70 for those older than age 62. Overall, this variable assessed the likelihood of working full-time in the next 5 years, on average. Second, responses were reported on a probability scale of 0 to 100 at the interval of 10 in KLoSA, whereas responses were on a continuous scale from 0 to 100 in the HRS. Finally, work expectations were assessed only among respondents who were working at the time of assessment in the KLoSA; in contrast, the HRS assessed work expectations in all respondents irrespective of their working status. This difference in the assessment of work expectations informed the basis of the sensitivity analyses described below.

In the main analysis, work expectations were modeled as a continuous time-varying exposure, and responses were rescaled to 0 to 10 for a more meaningful interpretation of regression coefficients (i.e., a one-unit increase in the scaled expectation is equivalent to a ten-unit increase in the original variable).

### Psychological distress

Psychological distress was assessed by the Center for Epidemiologic Studies Depression Scale (CES-D) on all respondents at every interview wave. The CES-D is originally a 20-item screening instrument that assesses four domains associated with distress symptoms (i.e., depressed effect, positive effect, somatic symptoms, and interpersonal relations) during the past week. Although the HRS and KLoSA both derive their psychological distress measures from the CES-D, the HRS uses an 8-item scale, each recorded as yes/no (CES-D 8; scores range from 0 to 8). In contrast, the KLoSA uses a 10-item scale, each recorded on a 4-point Likert scale ranging from “very rarely (less than 1 day)” to “almost always (5 to 7 days)” (CES-D 10; scores range from 0 to 30). Specific items in CES-D 8 and CES-D 10 are summarized in Supplementary Table 2. Previous studies have shown that CES-D 8 and CES-D 10 have comparable psychometric properties (O’Halloran et al., 2014), acceptable internal consistency, and test-retest reliability in older adults (Cronbach alpha is 0.78 – 0.84 for CES-D 8 and 0.72 – 0.80 for CES-D 10) (Cheng & Chan, 2005; Dang et al., 2020).

The CES-D summary scores were constructed by summing all symptoms, reverse-coding items on positive effects (e.g., “happiness” and “enjoying life”); higher scores indicated more severe symptomatology. To facilitate the comparison between the HRS and KLoSA, summary scores were then dichotomized to classify cases of psychological distress using threshold scores of ≥4 symptoms for CES-D 8 (HRS) and ≥10 for CES-D 10 (KLoSA). These thresholds have been shown to be equivalent to a threshold of 16 or more symptoms on the original 20-item CES-D (Steffick, 2000) and demonstrated acceptable sensitivity and specificity for distress symptoms in previous literature (Cheng & Chan, 2005; Dang et al., 2020).

### Employment transitions during the Recession

Types of employment transitions during the Recession were created in two steps. First, the employment status patterns were constructed for each respondent based on the labor force status reported in 2006, 2008, and 2010. Next, these employment patterns were manually classified into 6 employment transitions: entrance to full-time employment, precarious employment, stable full-time employment, self-employment or transition to self-employment, exit labor force, and disabled, retired, or being out of the labor force (detailed in Supplementary Table 3). These categories of employment transitions have been identified as common employment pathways during the Recession in previous literature (Kalousová & Burgard, 2022; J. Lee & Kim, 2017). They were time-invariant and reflected the employment trajectories the respondents experienced during the Recession.

### Covariates

All models adjusted for covariates related to demographics and health characteristics that have been associated with work expectations and psychological distress in prior work. Demographics included age (centered at each sample’s mean), sex (male, female), education (less than lower secondary education, upper secondary/vocational training, tertiary education), marital status (married, not married), minority status, household income, and total wealth. Minority status (yes/no) was indicated as racial/ethnic minority group for the US (e.g., non-Hispanic Black, Hispanics, and others) and rural residential status for Korea. Household income and total wealth were categorized into quartiles based on 2008 data. Health characteristics included self-rate health (poor/fair, good, very good/excellent) and medical morbidity, indicated by having at least one chronic condition among hypertension, diabetes, cancer, chronic lung disease, heart problem, stroke, and arthritis. All covariates were time-varying as a function of interview wave except for race/ethnicity and sex.

### Statistical approach

Initially, demographic and health characteristics at the 2008 wave were described for the full sample and by employment transitions, separately for the US and Korea.

Mixed effects logistic regressions with random intercept and random slope for age were used to examine the association between work expectations and psychological distress over the 12-year observation period. This modeling approach accounts for the clustered structure in the data and the time-varying nature of work expectations and psychological distress. It also incorporates information from all assessments rather than only those observations with complete follow-up over the study period (Hardin & Hilbe, 2018). The inclusion of random intercept term accounted for the heterogeneity in the baseline psychological distress; likelihood ratio test of the random intercept model vs. the model without was highly significant (p<0.0001). The random slope of age allowed each respondent to have a different rate of change in distress status over time.

Consistent with the official timeline of the Recession (Weinberg, 2013), indicators for different time periods were created based on the ending interview dates as follows: *pre*-*recession* (January 2006 – November 2007), *recession* (December 2007 – June 2009), and *recovery* (after June 2009). To examine how the Recession moderated the association between work expectations and psychological distress, these indicators and their interactions with work expectations were included in all models, using the recession period as reference. All analyses were performed separately for the US and Korea to reflect the differences in the socioeconomic contexts of the Recession in two settings.

Finally, stratification by employment transitions was performed to examine how employment experiences during the Recession moderated the association between work expectations and psychological distress. To visualize the findings, plots of average marginal predicted probabilities for psychological distress were displayed for the full sample and each employment transition.

Statistical analyses were performed in the R environment, version 4.2.2 (2022). Significance tests were evaluated two-sided at *p*<0.05. Mixed effects logistic regressions were fit via *glmer*, which estimates maximum likelihood by Laplace approximation. Marginal mean predicted probabilities were estimated with R package *ggeffects*, version 1.5.0.

### Sensitivity analyses

A series of sensitivity analyses were conducted to assess the robustness of the results. First, since there is no consensus on CES-D threshold scores, alternative thresholds (≥3, ≥5 symptoms for CES-D 8 and ≥11, ≥12 symptoms for CES-D 10) were used to classify cases of psychological distress (Cheng & Chan, 2005; Dang et al., 2020). Second, a set of analyses using the HRS was conducted that limited the measure of work expectations to those reported among people currently employed (at the time of assessment) to allow for a more comparable comparison with results from the KLoSA (KLoSA only assessed work expectations only among respondents who were working at the time of assessment). Finally, models were fit using the categorical versions of work expectations (operationalized as 0-40%, 41-60%, 61-89%, 90-100%) to explore potential non-linear associations between work expectations and psychological distress, with the category of 41-60% as the reference to approximate “50/50 chance”/uncertainty. These categories were constructed based on visual inspection of the histogram of work expectations.

## RESULTS

### Descriptive statistics

Table 1 shows the overall sample characteristics of the US and Korean respondents at the 2008 wave. In both countries, the majority were female, married, had upper secondary education or vocational training, were in good, very good, or excellent health, and were currently employed. Compared to the US, the Korean sample was younger (mean age: 50 vs. 57 years), had higher mean expectations of working in the next 5 years (75% vs. 37%), and lower prevalence of psychological distress (14% vs. 17%).

**Table 1.** Sample characteristics of United States and South Korean respondents in 2008

| **Sample characteristics in 2008** | **United States**<br>(N = 2,647) | **South Korea**<br>(N = 1,454) |
| --- | --- | --- |
| Number of interview waves (mean ± SD) | 5.9 ± 1.4 | 5.1 ± 1.9 |
| Total person-years | 25,725 | 12,096 |
| Psychological distress |  |  |
| Normalized CES-D score (mean ± SD) <sup>a</sup> | 0.20 ± 0.27 | 0.18 ± 0.14 |
| CES-D 8 ≥ 4 or CES-D 10 ≥ 10, N (%) | 436 (16.8) | 207 (14.3) |
| Work expectation |  |  |
| Mean ± SD | 37.1 ± 37.0 | 75.1 ± 23.0 |
| Median (interquartile range) | 25 (75) | 80 (40) |
| Age (mean ± SD) | 57.0 ± 1.8 | 49.9 ± 2.0 |
| Female, N (%) | 1,521 (57.5) | 745 (51.2) |
| Minority group, N (%) <sup>b</sup> | 847 (32.0) | 255 (17.5) |
| Education, N (%) |  |  |
| Lower secondary | 340 (12.8) | 448 (30.8) |
| Upper secondary or vocational training | 1,536 (58.0) | 780 (53.6) |
| Tertiary | 771 (29.2) | 226 (15.5) |
| Married or partnered, N (%) | 1,871 (70.7) | 1,322 (90.9) |
| Household income quartile, N (%) <sup>c</sup> |  |  |
| First (lowest) | 662 (25.0) | 364 (25.0) |
| Second | 663 (25.0) | 364 (25.0) |
| Third | 662 (25.0) | 386 (26.5) |
| Fourth (highest) | 660 (25.0) | 340 (23.4) |
| Total wealth quartile, N (%) <sup>d</sup> |  |  |
| First (poorest) | 662 (25.0) | 364 (25.1) |
| Second | 664 (25.1) | 363 (25.1) |
| Third | 660 (24.9) | 360 (24.9) |
| Fourth (richest) | 661 (25.0) | 361 (24.9) |
| Employment status, N (%) |  |  |
| Full-time | 1,285 (48.5) | 539 (37.1) |
| Part-time | 179 (6.8) | 104 (7.2) |
| Self-employed | 314 (11.9) | 465 (32.0) |
| Unpaid family work for 18+ hours <sup>e</sup> | - | 114 (7.8) |
| Unemployed | 81 (3.1) | 21 (1.4) |
| Partly retired/retired | 500 (18.9) | 61 (4.2) |
| Disabled or not in labor force | 288 (10.9) | 150 (10.3) |
| Self-rated health, N (%) |  |  |
| Poor/fair | 699 (26.4) | 384 (26.4) |
| Good | 760 (28.7) | 805 (55.4) |
| Very good/excellent | 1,188 (44.9) | 265 (18.2) |
| At least one chronic condition: hypertension, diabetes, cancer, chronic lung disease, heart problem, stroke, arthritis, N (%) | 1,920 (72.5) | 344 (23.7) |
| Current smoker, N (%) | 517 (19.5) | 363 (25.0) |
| Current drinker, N (%) | 1,072 (40.5) | 774 (53.2) |

<sup>a</sup> The normalized CES-D score for each respondent was calculated as the sum of all symptom scores divided by the maximum possible score, ranging from 0-1
<sup>b</sup> Minority status indicated a racial/ethnic minority group for the US (e.g., non-Hispanic Black, Hispanics, and others) and rural residential status for South Korea
<sup>c</sup> Quartiles based on household income of the analytical sample in 2008: US respondents (USD) <28,800, 28,800 - 61,999, 62,000 - 108,799, ≥108,800; South Korea respondents (10,000 Korean won) <1,620, 1,620 - 2,879, 2,880 - 4,235, ≥4,236
<sup>d</sup> Quartiles based on total wealth of the analytical sample in 2008: US respondents (USD) <32,000, 32,000 - 162,199, 162,200 - 482,499, ≥482,500; South Korea respondents (10,000 Korean won) <4,050, 4,050 - 10,316, 10,316 - 21,790, ≥21,790
<sup>e</sup> The employment category “unpaid family work for 18+ hours” was only in KLoSA

Between 2006 and 2010, approximately one-third of respondents were stably employed full-time, and less than one-fifth were in precarious employment (e.g., unemployed, temporary or part-time work) in both countries. While only 10% of US respondents were either self-employed or transitioning to self-employment during this period, almost one-third of Korean respondents underwent this employment transition. In the US, approximately 15% of respondents exited the labor force and 20% were retired or not in the labor force; these numbers in Korea were 7% and 5%, respectively (Supplementary Figure 2).

In the US, lowest work expectations were reported among respondents who exited the labor force (mean: 25.7, median: 10) or were retired/not in the labor force (mean: 5.4, median: 0). Respondents who were stably employed full-time had the lowest CES-D 8 score (mean: 1.0), whereas those who were not in the labor force had the highest score (mean: 2.8). Respondents who were stably employed full-time or self-employed were more likely to be male, non-Hispanic White, had at least upper secondary education, had higher household income and wealth, and reported very good or excellent health (Supplementary Table 4, p < 0.05). In Korea, similar results were found; however, the differences in work expectations among Korean respondents transitioning into different employment paths were relatively small than those observed among US respondents (Supplementary Table 5.5).

### Work expectations and psychological distress over the 12-year observation period

Figure 1 illustrates the association between work expectations and psychological distress, with a focus on the influence of the Recession. In general, higher expectations of working in the next 5 years were associated with lower odds of psychological distress after adjusting for demographics and health characteristics across all time periods (pre-recession, recession, and recovery). In the US, this inverse association was statistically significant during the Recession (odds ratio (OR) = 0.93, 95% confidence interval (CI): 0.89, 0.98 for every 10% increase in work expectations) and recovery periods (OR = 0.95 [95%CI: 0.92, 0.98]). In Korea, the inverse association between work expectations and psychological distress was more pronounced than in the US, particularly during the recovery (OR = 0.84 [95%CI: 0.80, 0.89]).

**Figure 1.**
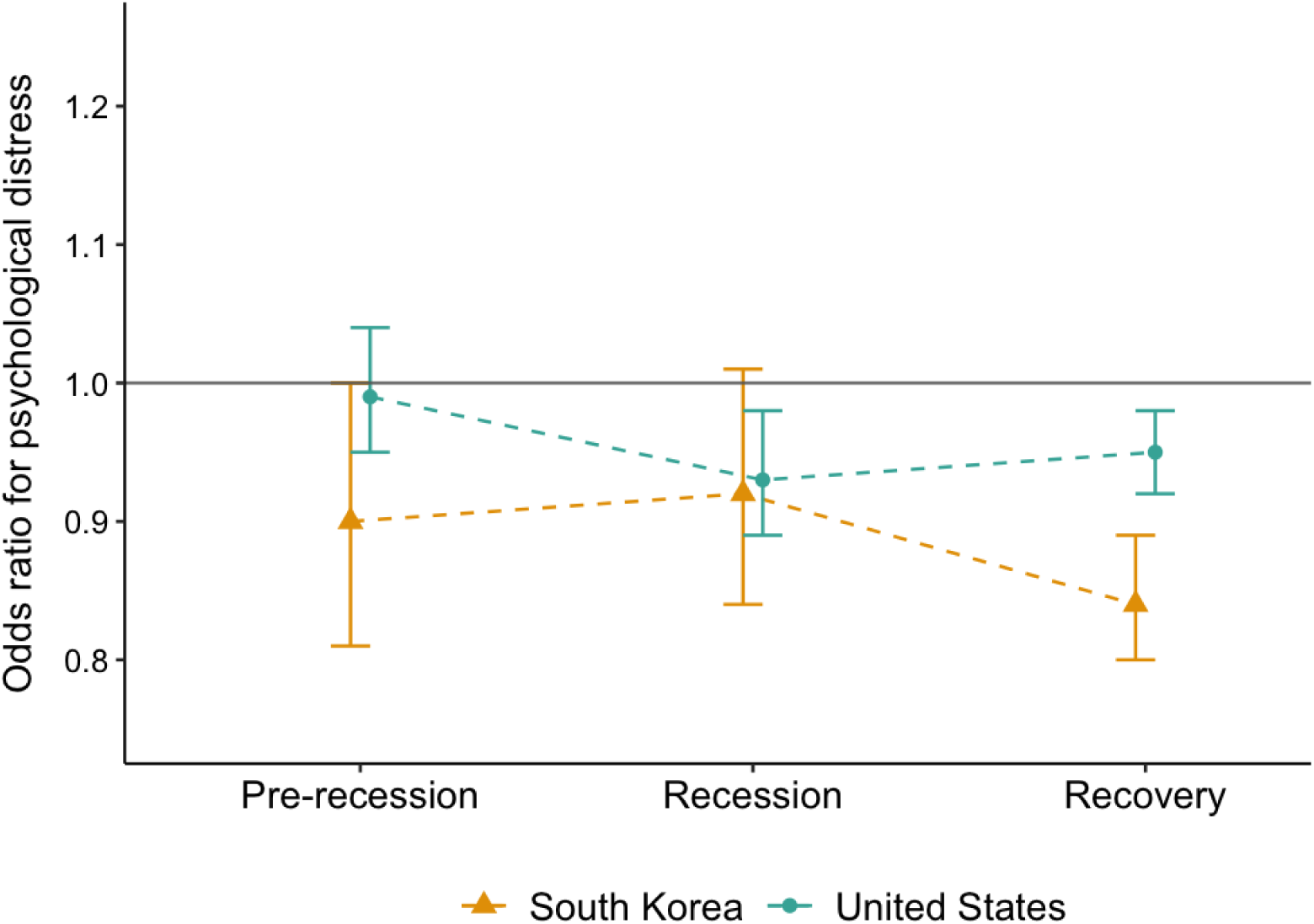
Adjusted odds ratios for the association between work expectations and psychological distress in the United States and South Korea from 2006 to 2018 Note: ^a^ Odds ratios for having CES-D 8 ≥ 4 (United States) or CES-D 10 ≥ 10 (South Korea) were estimated on the full analytical samples; 95% confidence intervals were approximated using the delta method. Work expectations were modeled as a continuous variable and scaled to 0-10. ^b^ All models adjusted for centered age, sex, minority status, education, marital status, household income, total wealth, self-rated health, having at least one chronic condition, random intercept, and random slope for age ^c^ Three time periods of the Recession were created based on the ending interview dates according to the official timeline of the Recession (Weinberg, 2013): pre-recession period (before December 2007), recession period (December 2007 - June 2009), and recovery period (after June 2009)

### Work expectations and psychological distress by employment transitions

Table 2 shows how employment transitions moderated the association between work expectations and psychological distress during three time periods of the Recession. In the US, for those who were stably employed full-time or precariously employed, work expectations were not associated with psychological distress. Similarly, there were no associations among those who exited the labor force or were disabled, retired, or not in the labor force. However, there was a robust negative influence of the Recession among respondents who transitioned into self- or full-time employment. Specifically, during the Recession period, higher expectations were associated with higher odds of distress among self-employed respondents (OR = 1.25 [95% CI: 1.03, 1.52]), but lower odds among those who transitioned into full-time employment (OR = 0.74 [95% CI: 0.57, 0.98]).

**Table 2.** Adjusted odd ratios and 95% confidence intervals for the association between work expectations and psychological distress by employment transition in the United States and South Korea over a 12-year period

|  | Odds ratio (95% confidence interval) |  |  |
| --- | --- | --- | --- |
|  | Pre-recession<br>(2006 - Nov 2007) | Recession<br>(Dec 2007 - Jun 2009) | Recovery<br>(July 2009 - 2018) |
| **United States** |  |  |  |
| Full sample (N = 2,647) | 0.99 (0.95, 1.04) | 0.93 (0.89, 0.98)* | 0.95 (0.92, 0.98)* |
| Entrance to full-time (N = 122) | 1.31 (1.04, 1.65)* | 0.74 (0.57, 0.98)* | 1.00 (0.86, 1.16) |
| Precarious employment (N = 361) | 0.99 (0.88, 1.11) | 0.97 (0.86, 1.09) | 0.95 (0.87, 1.04) |
| Stable full-time (N = 884) | 0.98 (0.88, 1.09) | 1.00 (0.90, 1.13) | 0.95 (0.89, 1.02) |
| Self-employment, transition to self-employment (N = 275) | 1.09 (0.93, 1.29) | 1.25 (1.03, 1.52)* | 1.04 (0.93, 1.15) |
| Exit labor force (N = 414) | 0.99 (0.89, 1.09) | 0.90 (0.80, 1.02) | 0.97 (0.87, 1.08) |
| Disabled, retired, out of labor force (N = 591) | 0.98 (0.85, 1.13) | 1.05 (0.87, 1.26) | 0.92 (0.80, 1.07) |
| **South Korea** |  |  |  |
| Full sample (N = 1,454) | 0.90 (0.81, 1.00)* | 0.92 (0.84, 1.01) | 0.84 (0.80, 0.89)* |
| Entrance to full-time (N = 117) | 0.69 (0.32, 1.50) | 1.01 (0.74, 1.37) | 0.97 (0.78, 1.22) |
| Precarious employment (N = 269) | 0.88 (0.70, 1.11) | 0.91 (0.74, 1.12) | 0.88 (0.79, 0.99)* |
| Stable full-time (N = 418) | 0.95 (0.78, 1.16) | 0.95 (0.79, 1.14) | 0.91 (0.83, 1.00)* |
| Self-employment, transition to self-employment (N = 432) | 0.86 (0.70, 1.05) | 0.86 (0.73, 1.01) | 0.75 (0.68, 0.83)* |
| Exit labor force (N = 122) | 0.89 (0.50, 1.58) | 1.19 (0.66, 2.16) | 0.60 (0.30, 1.18) |

<sup>a</sup> Odds ratios were calculated for having CES-D 8 ≥ 4 (United States) or CES-D 10 ≥ 10 (South Korea); 95% confidence intervals were approximated using the delta method. Work expectations were modeled as a continuous variable and scaled to 0-10
<sup>b</sup> All models adjusted for centered age, sex, minority status, education, marital status, household income, total wealth, self-rated health, having at least one chronic condition, random intercept, and random slope for age
<sup>c</sup> Model did not converge among South Korean respondents who were retired or not in the labor force
\* $p < .05$

In Korea, there were no associations between work expectations and psychological distress among respondents who transitioned into full-time employment or exited the labor force. Similar to the US, the effect of the Recession was also robustly observed among self-employed respondents; however, the association was in the opposite direction compared to the US. Specifically, higher expectations were associated with lower odds of psychological distress among self-employed respondents, a relationship that was statistically significant during the recovery period (OR = 0.75 [0.68, 0.83]).

Figure 2 shows the average marginal predicted probability of psychological distress associated with work expectations during the Recession and recovery periods in the US and Korea. In both countries, higher work expectations were inversely associated with psychological distress overall. In Korea, during the Recession, higher expectations were inversely associated with psychological distress among respondents who were self-employed, precariously employed, or stably full-time employed (Figure 2A). This inverse relationship was observed among all employment transitions during the recovery period (Figure 2B). In the US, during the Recession, higher work expectations were positively associated with psychological distress among self-employed respondents; in contrast, higher expectations were inversely associated with distress among those who transitioned into full-time employment (Figure 2C). During the recovery period, there was a weak inverse association between higher expectations and lower predicted probabilities of psychological distress overall (Figure 2D).

**Figure 2.**
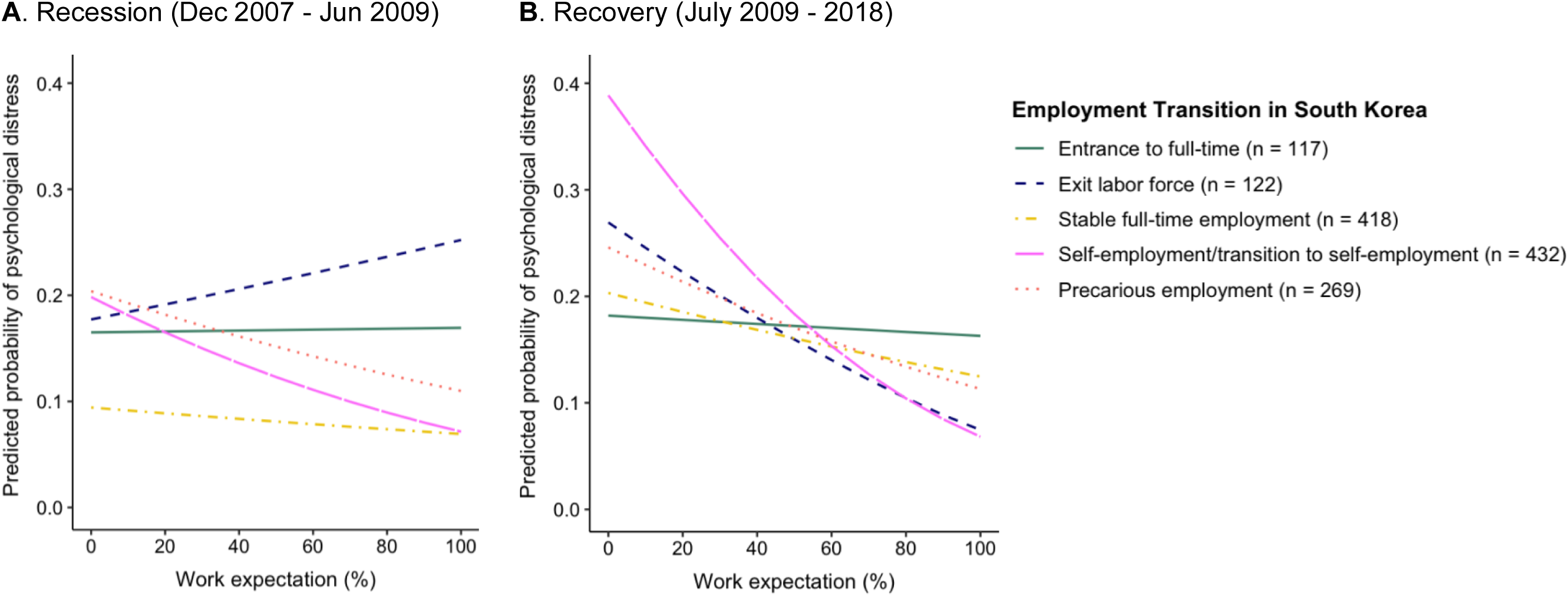
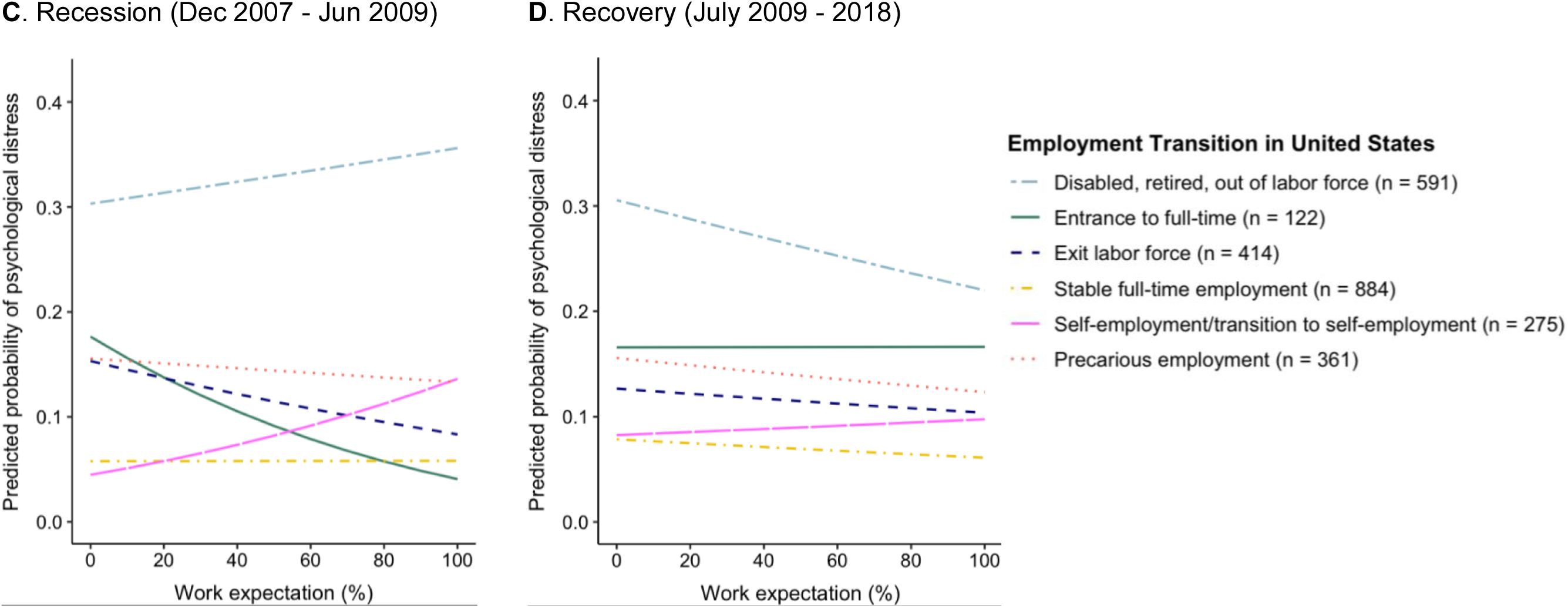
Average marginal predicted probability of psychological distress by employment transition in the United States and South Korea during the Great Recession and its recovery Note: ^a^ Average marginal predicted probabilities for having CES-D 8 ≥ 4 (United States) or CES-D 10 ≥ 10 (South Korea) were estimated using continuous work expectations (measured on a scale of 0-100%) ^b^ All models adjusted for centered age, sex, minority status, education, marital status, household income, total wealth, self-rated health, having at least one chronic condition, random intercept and random slope for age ^c^ Time periods of the Recession were created based on the ending interview dates according to the official timeline of the Recession (Weinberg, 2013): recession period (December 2007 - June 2009) and recovery period (after June 2009)

### Sensitivity analyses

Results were similar when using alternative threshold scores of CES-D for classifying psychological distress (Supplementary Table 6 and 7). When analyses in the HRS were limited to measures of expectations reported while the respondent was currently employed (to better align with the assessment of this construct in KLoSA), results were also similar (Supplementary Table 8). Finally, analyses using the categorical version of work expectations yielded similar results, though small sample sizes affected the precision of estimates (Supplementary Table 9 and 10).

## DISCUSSION

This study examined the association between work expectations and psychological distress and how employment transitions moderated this association during the Recession among the US and Korean Baby Boomers. There are four major findings: First, higher expectations of working in the next 5 years were inversely associated with psychological distress over the 12-year period that spanned the Recession. Second, this inverse association was moderated by the macroeconomic context, such that it was most robustly observed during the recession and recovery periods. Third, the association between expectations and psychological distress varied by employment transitions and was particularly pronounced among respondents who were self-employed or transitioning into self-employment. Finally, the relationships between work expectations, employment transitions, and psychological distress were broadly similar, with some notable differences, comparing the US and Korea.

As hypothesized, the inverse association between work expectations and psychological distress was particularly robust during the recession and recovery periods. This finding is consistent with theories of cumulative stress and personal control on the causes of psychological distress (Koltai & Stuckler, 2019). Previous literature has linked psychological distress to the cumulative experience of multiple stressors not only through economic hardships (e.g., job loss, financial strain) (Tsuchiya et al., 2020) but to a greater extent, perceived job and employment insecurity relating to the Recession (Burgard et al., 2009). Concerns about job insecurity (i.e., anticipation of involuntarily losing current job) and reemployment insecurity (i.e., likelihood of reemployment in the event of job loss) inherently evoke perceptions of the future. Both constructs are tied to individual expectations of future employment prospects and cognitive coping such as sense of personal control (Frone, 2018). Expecting to be working in the future may reflect higher levels of personal control despite an uncertain economic climate, which in turn protects against adverse mental health consequences during recessions (Koltai & Stuckler, 2019).

Furthermore, the study demonstrates that heterogeneity in employment transitions is an important moderator of the association between the Recession and psychological distress among older workers. This finding may reflect self-selection into employment pathways, differential stress exposures, or differences in coping resources. Theories related to cumulative inequality (Crystal et al., 2017) and the “healthy worker” effect (Li & Sung, 1999) posit that demographic and health characteristics are the main drivers of self-selection into a particular employment path. The present study provides empirical evidence for these theories in two ways. First, it shows that respondents with stable full-time employment had higher socioeconomic status and better health than those in other employment transitions. Second, those in unfavorable employment paths (e.g., being precariously employed, not in the labor force) had lower expectations for future work and higher distress symptoms. Moreover, social and health determinants are closely tied to the theory of differential stress exposures. Drawing on this theory, study findings suggest that individuals with disadvantaged social and health status are at a greater risk for negative employment experiences and consequently more distress symptoms. Finally, variation in the relationships between work expectations, employment transitions and psychological distress may reflect individual differences in coping responses to stressful economic environments. Those in unfavorable employment paths may experience heightened feelings of uncertainty (Frone, 2018) and hopelessness (Parada-Fernández et al., 2020), which then prompts maladaptive coping responses and elevated distress symptoms (Pearlin & Bierman, 2013).

Results from the cross-country comparison between the US and Korea are two-fold. First, employment transitions varied between two countries. In particular, a higher proportion of US respondents exited or were not in the labor force during the Recession versus Korea. This finding may reflect the difference in severity of the Recession between the two countries. In the US, the unemployment rate increased significantly during this period. In contrast, Korea experienced minimal impact from the Recession, maintaining a relatively stable unemployment rate throughout the crisis (Arias & Wen, 2015). Moreover, American respondents were older (mean age of 57 years), so they may have been more likely to retire or exit the labor force when the prospect of finding stable employment became challenging. Second, the moderating effect of employment transitions on the association between work expectations and psychological distress varied between countries. Notably, higher expectations were associated with higher odds of distress among self-employed American adults, but lower odds of distress among self-employed Korean adults. This discrepancy should be interpreted in consideration of the impact of the Recession on workforce characteristics. The severity of the Recession increased the number of workers who transitioned into self-employment as a means to avoid prolonged unemployment and financial hardship (Beckhusen, 2014a). Research on self-employment and psychological distress is mixed, with some studies reporting that self-employed workers experienced less distress, while others finding null or positive associations between self-employment and distress (Willeke et al., 2021). These inconsistent findings may reflect that in some situations self-employment is a voluntary choice, whereas in others it is an involuntary transition (Park et al., 2022). In Korea, many older workers involuntarily retire early from their full-time employment because of the widespread practice of honorary retirement (OECD, 2018). It is possible that these workers may view self-employment as an expected career transition following retirement from their “main” jobs, and thus are more inclined to choose self-employment as a financial means to earn their living during the economic crisis. The net result is that higher expectations of continuing self-employment may bring psychological benefits for older Korean workers. On the contrary, given the more severe impact of the Recession in the US, workers may have more difficulty in starting or retaining their businesses and thus experienced worse psychological distress.

### Implications for policies regarding work and retirement

Overall, the study highlights the inequality in employment experiences and its implications for psychological distress among older workers. Beyond employment status, workplace policies should consider work transitions (e.g., expected or unexpected) as well as the preferences and constraints that may drive these transitions. Effective policies may target communication strategies to better understand workers’ needs, offer flexibility in terms of work and schedules, and provide financial wellness resources and matched savings programs, regardless of employment status, to ease workers’ financial concerns during economic recession. These intervention strategies are particularly relevant in light of recent economic crisis and rising inflation brought by the COVID-19 pandemic. While the COVID-19 recession originated from different causes than the Recession, the economic consequences of the two recessions are fairly similar in many aspects, including shrinking economic activities and rising unemployment and inflation. Future studies may expand the present analysis to examine the impacts of the COVID-19 recession on employment expectations and psychological distress. Comparative studies between two recessions can help identify similar or distinctive risk and protective factors and potential mechanisms underlying the recession-related psychological distress among older workers transitioning into retirement and other employment paths.

### Strengths and limitations

Findings should be interpreted considering study limitations and strengths. First, there could be misclassification of employment transitions because they were determined based on employment statuses reported from 2006 to 2010. However, the shares of respondents undergoing each employment transition are consistent with the estimates reported from national surveys or census data for each country. Further, the study did not have available data to clarify the motivation and constraints that drive their selection into a particular employment path. Such data can shed insights into the underlying mechanisms linking employment transitions (e.g., expectedness, voluntariness) to psychological distress. Second, possible reverse causation may confound the results given that psychological distress could affect the expectations and ability to work. Future studies could address the potential of reverse causality by including data on early life experiences and prior diagnoses of psychological distress. Third, the analytical sample was limited to respondents with available data during the Recession; these respondents were on average older and more likely to be working. Thus, findings may not extend to younger older adults or those who were not currently working but look for jobs or re-enter the workforce. Finally, psychological distress was assessed by the CES-D rather than clinical interviews/diagnoses. Future studies could use more rigorous depression measures as well as examine other mental health outcomes (e.g., anxiety).

Despite these limitations, this study has several strengths. The study contributes to existing literature by providing empirical evidence for the adverse effects of the Recession on employment and psychological distress in older adults. While most studies on the Recession have been conducted in Western countries, this study situated the relationship between work expectations and psychological distress within the US and Korea. This cross-cultural comparison provides important insights into how the economic contexts and policy responses to the Recession of a given country influence individuals’ decisions about work and its implications for psychological distress. Future research can expand this analysis to other countries to gain a more comprehensive, international perspective of how macroeconomic events moderate the relationship between work expectations and psychological distress. Insights from such comparisons help to inform culturally appropriate interventions to support the mental health needs of an increasingly diverse population during the economic crisis. Furthermore, analyses on employment transitions provide more nuanced insights into workers’ diverse employment experiences during the Recession and its implications for psychological distress. Results were also robust across several sensitivity analyses. Finally, the study sample came from large population-based studies with rich data on employment and distress measures, which enhances the study’s validity.

## CONCLUSION

The association between work expectations and psychological distress was most robustly observed during the recession and recovery periods. Findings highlight the moderating roles of employment transitions and country contexts in shaping employment preferences and psychological distress. Cross-country comparative studies can help identify potential mechanisms underlying the recession-related psychological distress among older workers transitioning into retirement and other employment paths.

## Data Availability

All data are available online at https://g2aging.org.

## ACKNOWLEDGMENTS

The HRS and KLoSA are publicly available to researchers at https://hrs.isr.umich.edu and https://survey.keis.or.kr/eng/klosa/klosa01.jsp.

## TABLES/FIGURES

**Supplementary Figure 1.**
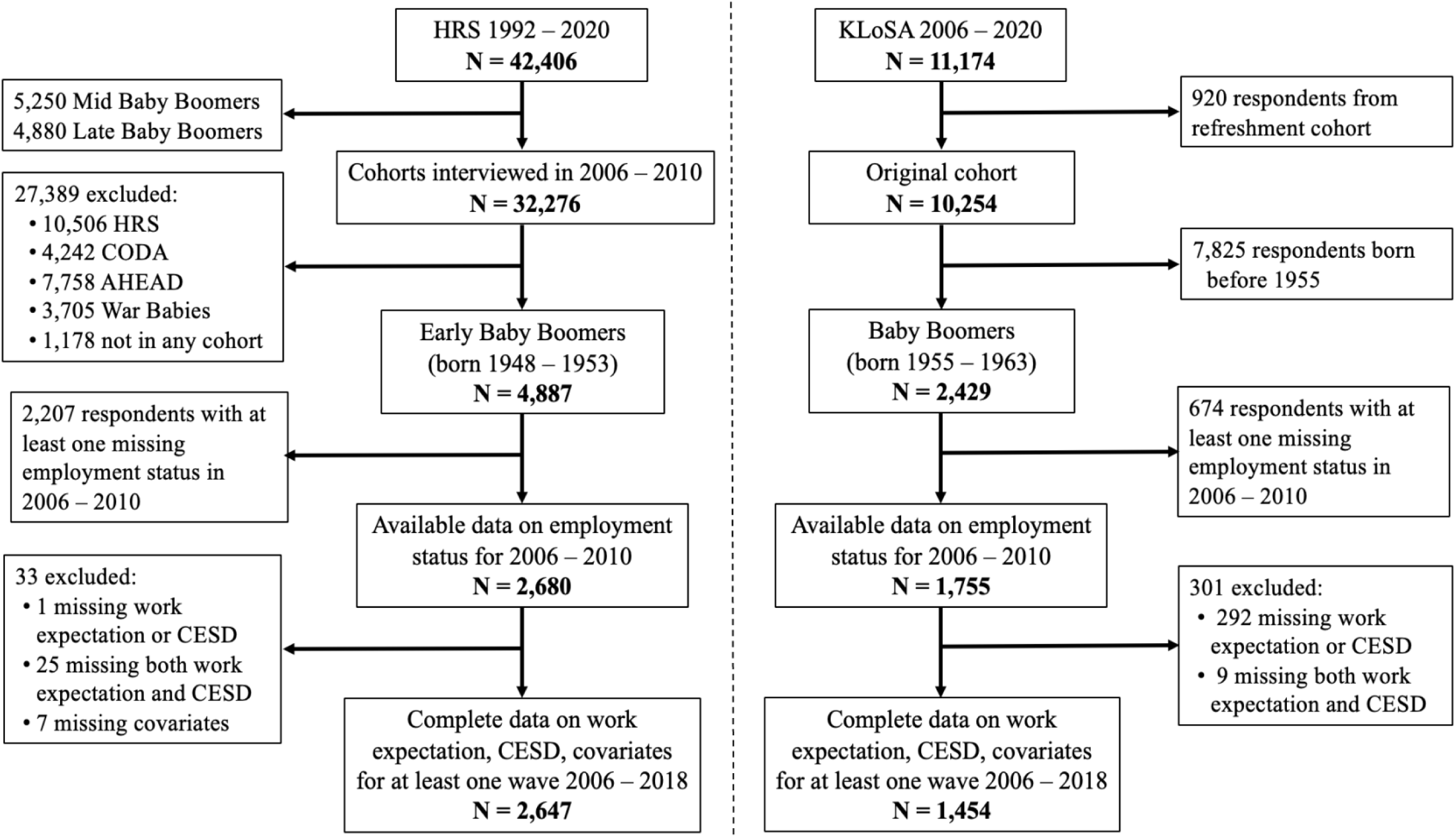
Selection criteria of the analytical samples in the Health and Retirement Study and Korean Longitudinal Study of Aging **Abbreviations**: HRS, Health and Retirement Study; KLoSA, Korean Longitudinal Study of Aging; CESD, Center for Epidemiologic Studies Depression Scale; CODA, Children of the Depression; AHEAD, Asset and Health Dynamics Among the Oldest Old

**Supplementary Table 1.** Sample characteristics of the analytical sample and excluded respondents at the 2008 interview wave in the United States and South Korea

| Sample characteristics in 2008 | United States |  |  | South Korea |  |  |
| --- | --- | --- | --- | --- | --- | --- |
|  | Included<br>(N = 2,647) | Excluded<sup>a</sup><br>(N = 14,570) | p-value<sup>b</sup> | Included<br>(N = 1,454) | Excluded<sup>a</sup><br>(N = 7,234) | p-value<sup>b</sup> |
| Normalized CES-D<br>(mean ± SD)<sup>c</sup> | 0.20 ± 0.27 | 0.18 ± 0.24 | 0.0004 | 0.18 ± 0.14 | 0.26 ± 0.19 | < .0001 |
| Age (mean ± SD) | 57.0 ± 1.8 | 71.4 ± 10.2 | < .0001 | 49.9 ± 2.0 | 66.4 ± 9.8 | < .0001 |
| Female, N (%) | 1,521 (57.5) | 8,672 (59.5) | 0.05 | 745 (51.2) | 4,177 (57.7) | < .0001 |
| Minority group, N (%)<sup>d</sup> | 847 (32.0) | 3,644 (25.0) | < .0001 | 255 (17.5) | 1,884 (26.0) | < .0001 |
| Education, N (%) |  |  |  |  |  |  |
| Lower secondary | 340 (12.8) | 3,393 (23.3) |  | 448 (30.8) | 5,014 (69.3) |  |
| Upper secondary,<br>vocational training | 1,536 (58.0) | 8,315 (57.1) | < .0001 | 780 (53.6) | 1,648 (22.8) | < .0001 |
| Tertiary | 771 (29.2) | 2,858 (19.6) |  | 226 (15.5) | 572 (7.9) |  |
| Married or partnered, N (%) | 1,871 (70.7) | 9,140 (62.7) | < .0001 | 1,322 (90.9) | 5,389 (74.5) | < .0001 |
| Household income quartiles, N(%)<sup>e</sup> |  |  |  |  |  |  |
| First (lowest) | 662 (25.0) | 5,995 (41.1) |  | 364 (25.0) | 4,953 (68.6) |  |
| Second | 663 (25.0) | 4,543 (31.2) |  | 364 (25.0) | 1,154 (16.0) |  |
| Third | 662 (25.0) | 2,251 (15.4) | < .0001 | 386 (26.5) | 574 (7.9) | < .0001 |
| Fourth (highest) | 660 (25.0) | 1,781 (12.2) |  | 340 (23.4) | 542 (7.5) |  |

| Total wealth quartiles, N(%) <sup>f</sup> |  |  |  |  |  |  |
| --- | --- | --- | --- | --- | --- | --- |
| First (poorest) | 662 (25.0) | 3,092 (21.2) |  | 364 (25.2) | 2,527 (35.0) |  |
| Second | 664 (25.1) | 3,414 (23.4) | < .0001 | 358 (24.8) | 1,740 (24.1) | < .0001 |
| Third | 660 (24.9) | 3,898 (26.8) |  | 364 (25.2) | 1,493 (20.7) |  |
| Fourth (richest) | 661 (25.0) | 4,166 (28.6) |  | 358 (24.8) | 1,459 (20.2) |  |
| Self-rated health |  |  |  |  |  |  |
| Poor/fair | 699 (26.4) | 4,559 (31.3) |  | 384 (26.4) | 4,326 (59.8) |  |
| Good | 760 (28.7) | 4,754 (32.7) | < .0001 | 805 (55.4) | 2,394 (33.1) | < .0001 |
| Very good/excellent | 1,188 (44.9) | 5,243 (36.0) |  | 265 (18.2) | 514 (7.1) |  |
| At least one: hypertension, diabetes, cancer, chronic lung disease, heart problem, stroke, arthritis, N(%) | 1,920 (72.5) | 12,818 (88.0) | < .0001 | 344 (23.7) | 4,230 (58.5) | < .0001 |
| Current smoker, N(%) | 517 (19.5) | 1,717 (11.9) | < .0001 | 363 (25.0) | 1,219 (16.9) | < .0001 |
| Current drinker, N(%) | 1,072 (40.5) | 4,538 (31.2) | < .0001 | 774 (53.2) | 2,386 (33.0) | < .0001 |

<sup>a</sup> Not all respondents were interviewed in 2008 (reasons: loss of follow-up, death, too young to be interviewed)
<sup>b</sup> Wilcoxon rank sum tests (comparing means) and Kruskal-Wallis tests (comparing medians) were used for continuous variables; chi-squared tests were used for categorical variables
<sup>c</sup> The normalized CES-D score for each respondent was calculated as the sum of all symptom scores divided by the maximum possible score, ranging from 0-1
<sup>d</sup> Minority status indicated a racial/ethnic minority group for the US (e.g., non-Hispanic Black, Hispanics, and others) and rural residential status for South Korea
<sup>e</sup> Quartiles based on household income of the analytical sample in 2008: US respondents (USD) <28,800, 28,800 - 61,999, 62,000 - 108,799, ≥108,800; South Korea respondents (10,000 Korean won) <1,620, 1,620 - 2,879, 2,880 - 4,235, ≥4,236
<sup>f</sup> Quartiles based on total wealth of the analytical sample in 2008: US respondents (USD) <32,000, 32,000 - 162,199, 162,200 - 482,499, ≥482,500; South Korea respondents (10,000 Korean won) <4,050, 4,050 - 10,316, 10,316 - 21,790, ≥21,790

**Supplementary Table 2.**
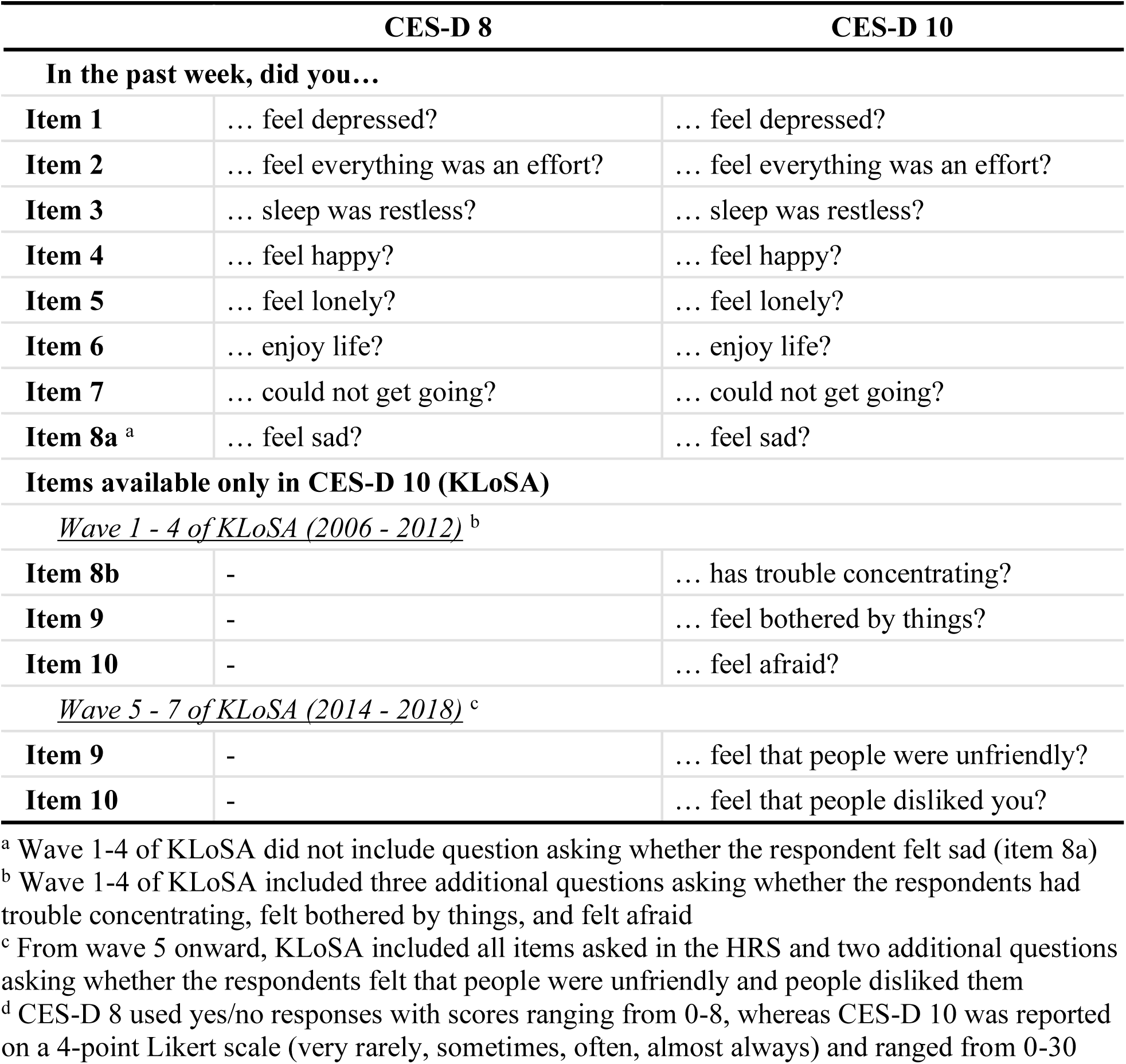
Items included in the 8-item (Health and Retirement Study, HRS) and 10-item (Korean Longitudinal Study of Aging, KLoSA) Center for Epidemiologic Studies – Depression Scale

**Supplementary Table 3.** Definitions of 6 employment transitions during the Great Recession

| Employment transition | Description |
| --- | --- |
| Entrance to full-time employment | Transition to full-time employment after periods of not working (e.g., unemployed, retired, not in the labor force), being previously self-employed, or having only part-time employment |
| Precarious employment | • Unemployment, part-time employment, or doing unpaid family work for 2+ interview waves 2006 – 2010<br>• Frequently changing employment status, i.e., switching between full-time, part-time, self-employment, unemployment, unpaid family work and periods of not working (e.g., retired)<br>• Employed with discontinuity (e.g., working full-time in 2006 and 2010 but unemployed in 2008)<br>• Under-employment (e.g., employed full-time in 2006 but become part-time or unemployed during the Recession) |
| Stable full-time employment | Full-time employment in all waves 2006 – 2010 |
| Self-employment, transition to self-employment | • Self-employment in all waves 2006 – 2010<br>• Transition to self-employment from previous full/part-time employment or periods of not working (e.g., unemployed, retired, not in labor force) |
| Exit labor force | • Transition out of the labor force from being unemployed during the Recession (2006 – 2008) or from full/part-time employment or self-employment<br>• Reducing to part-time employment or partly retirement from full-time or self-employment<br>• Exit labor force due to disability |
| Disabled, retired, or out of labor force | Not in the labor force (e.g., retired, on disability) for 2+ interview waves 2006 – 2010 |

**Supplementary Figure 2.**
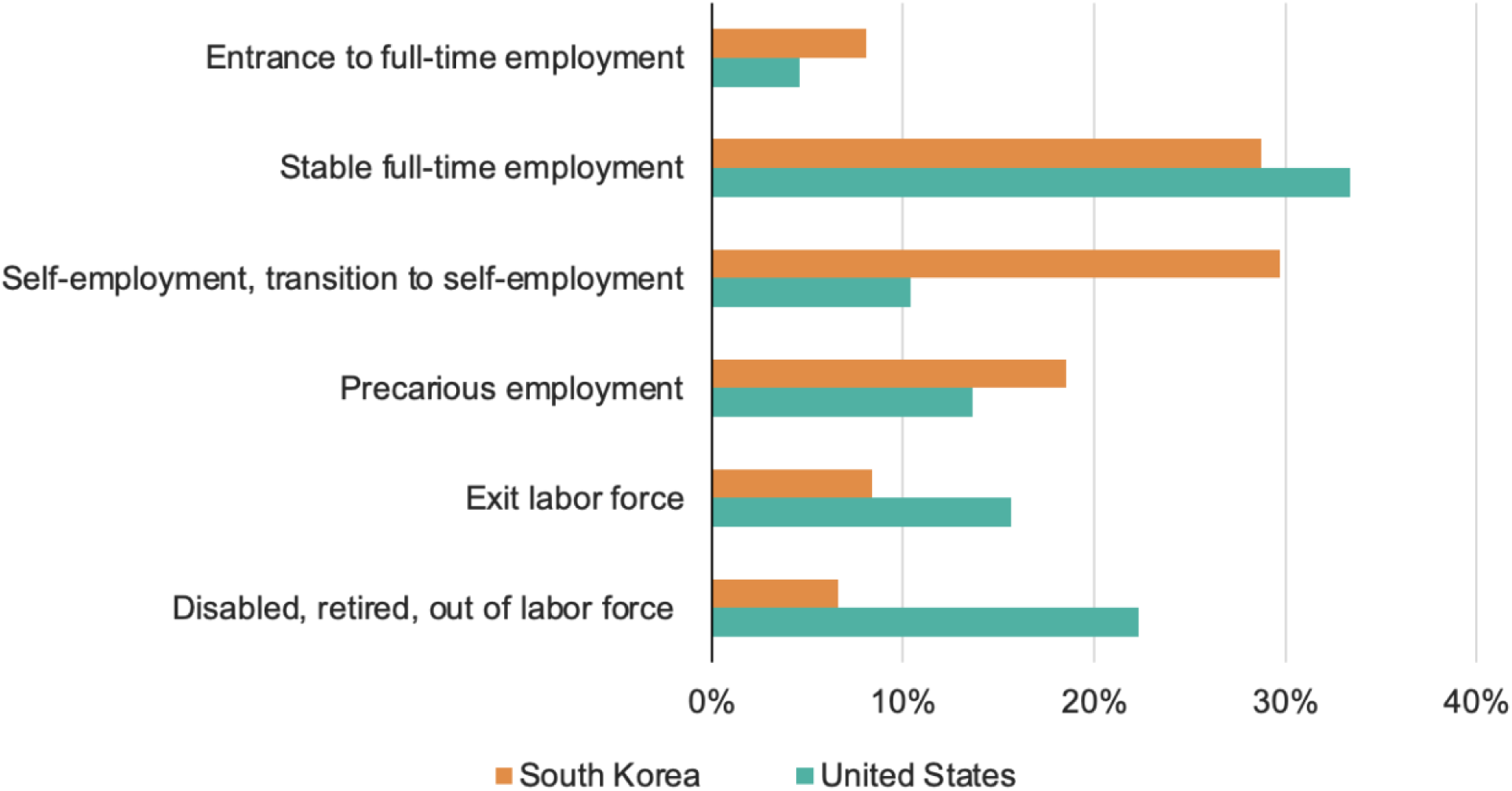
Employment transitions in the United States and South Korea between 2006 to 2010

**Supplementary Table 4.** Sample characteristics at the 2008 wave for US respondents by employment transitions experienced during the Recession

| Sample characteristics in 2008 | Employment transition during the Great Recession |  |  |  |  |  |
| --- | --- | --- | --- | --- | --- | --- |
|  | Entrance to full-time | Precarious employment | Stable full-time | Self-employment or transition to self-employment | Exit labor force | Disabled, retired, out of labor force |
|  | (N = 122) | (N = 361) | (N = 884) | (N = 275) | (N = 414) | (N = 591) |
| Number of interview waves (mean ± SD) | 6.0 ± 1.2 | 5.9 ± 1.3 | 5.9 ± 1.3 | 6.0 ± 1.2 | 5.8 ± 1.4 | 5.6 ± 1.5 |
| Total person-years | 1,216 | 3,564 | 8,731 | 2,745 | 4,005 | 5,464 |
| Psychological distress |  |  |  |  |  |  |
| CES-D 8 score (max:8; mean ± SD) | 1.3 ± 1.9 | 1.7 ± 2.2 | 1.0 ± 1.6 | 1.1 ± 1.7 | 1.5 ± 2.0 | 2.8 ± 2.7 |
| CES-D 8 ≥ 3, N (%) | 24 (20.0) | 93 (26.3) | 97 (11.2) | 42 (15.4) | 88 (21.7) | 255 (44.3) |
| CES-D 8 ≥ 4, N (%) | 17 (14.2) | 68 (19.3) | 65 (7.5) | 24 (8.8) | 61 (15.1) | 201 (35.0) |
| CES-D 8 ≥ 5, N (%) | 8 (6.7) | 50 (14.2) | 46 (5.3) | 18 (6.6) | 40 (9.9) | 160 (27.8) |
| Work expectation |  |  |  |  |  |  |
| Mean ± SD | 46.6 ± 34.5 | 41.6 ± 35.4 | 53.6 ± 36.3 | 56.4 ± 34.1 | 25.7 ± 31.8 | 5.4 ± 15.2 |
| Median (interquartile range) | 50 (70) | 40 (70) | 50 (70) | 60 (60) | 10 (50) | 0 (0) |
| Age (mean ± SD) | 56.9 ± 1.7 | 56.9 ± 1.8 | 56.8 ± 1.7 | 56.8 ± 1.8 | 57.4 ± 1.8 | 57.2 ± 1.8 |
| Female, N (%) | 81 (66.4) | 234 (64.8) | 433 (49.0) | 127 (46.2) | 244 (58.9) | 402 (68.0) |
| Racial/ethnic minority, N (%) | 41 (33.6) | 128 (35.5) | 240 (27.1) | 65 (23.6) | 130 (31.4) | 243 (41.1) |
| Education, N(%) |  |  |  |  |  |  |
| Lower secondary | 17 (13.9) | 48 (13.3) | 62 (7.0) | 20 (7.3) | 42 (10.1) | 151 (25.5) |
| Upper secondary, vocational training | 65 (53.3) | 219 (60.7) | 517 (58.5) | 139 (50.5) | 254 (61.4) | 342 (57.9) |
| Tertiary | 40 (32.8) | 94 (26.0) | 305 (34.5) | 116 (42.2) | 118 (28.5) | 98 (16.6) |
| Married/partnered, N (%) | 85 (69.7) | 260 (72.0) | 646 (73.2) | 218 (79.3) | 288 (69.6) | 374 (63.3) |
| Household income (in USD), N(%) |  |  |  |  |  |  |
| < 28,800 | 28 (23.0) | 97 (26.9) | 75 (8.5) | 37 (13.5) | 104 (25.1) | 321 (54.3) |
| 28,800 - 61,999 | 34 (27.9) | 100 (27.7) | 254 (28.7) | 50 (18.2) | 97 (23.4) | 128 (21.7) |
| 62,000 - 108,799 | 33 (27.0) | 84 (23.3) | 293 (33.1) | 71 (25.8) | 95 (22.9) | 86 (14.6) |
| ≥ 108,800 | 27 (22.1) | 80 (22.2) | 262 (29.6) | 117 (42.5) | 118 (28.5) | 56 (9.5) |
| Total wealth (in USD), N(%) |  |  |  |  |  |  |
| < 32,000 | 29 (23.8) | 95 (26.3) | 165 (18.7) | 31 (11.3) | 99 (23.9) | 243 (41.1) |
| 32,000 - 162,199 | 32 (26.2) | 91 (25.2) | 279 (31.6) | 48 (17.5) | 85 (20.5) | 129 (21.8) |
| 162,200 - 482,499 | 33 (27.0) | 94 (26.0) | 249 (28.2) | 79 (28.7) | 105 (25.4) | 100 (16.9) |
| ≥ 482,500 | 28 (23.0) | 81 (22.4) | 191 (21.6) | 117 (42.5) | 125 (30.2) | 119 (20.1) |
| Self-rated health |  |  |  |  |  |  |
| Poor/fair | 26 (21.3) | 78 (21.6) | 119 (13.5) | 43 (15.6) | 115 (27.8) | 318 (53.8) |
| Good | 42 (34.4) | 98 (27.1) | 294 (33.3) | 64 (23.3) | 130 (31.4) | 132 (22.3) |
| Very good/excellent | 54 (44.3) | 185 (51.2) | 471 (53.3) | 168 (61.1) | 169 (40.8) | 141 (23.9) |
| At least one chronic condition:<br>hypertension, diabetes, cancer, chronic<br>lung disease, heart problem, stroke,<br>arthritis, N(%) | 83 (68.0) | 241 (66.8) | 614 (69.5) | 176 (64.0) | 307 (74.2) | 499 (84.4) |
| Current smoker, N(%) | 24 (19.7) | 76 (21.1) | 136 (15.4) | 41 (14.9) | 71 (17.1) | 169 (28.6) |
| Current drinker, N(%) | 49 (40.5) | 156 (43.2) | 398 (45) | 141 (51.3) | 175 (42.3) | 153 (25.9) |

**Supplementary Table 5.** Sample characteristics at the 2008 wave for Korean respondents by employment transitions experienced during the Recession

| Sample characteristics<br>in 2008 | Employment transition during the Great Recession |  |  |  |  |  |
| --- | --- | --- | --- | --- | --- | --- |
|  | Entrance to<br>full-time | Precarious<br>employment | Stable<br>full-time | Self-employment<br>or transition to<br>self-employment | Exit labor<br>force | Disabled,<br>retired, out of<br>labor force |
|  | (N = 117) | (N = 269) | (N = 418) | (N = 432) | (N = 122) | (N = 96) |
| Number of interview wave (mean ± SD) | 4.6 ± 1.8 | 5.2 ± 1.7 | 5.8 ± 1.4 | 5.9 ± 1.5 | 2.9 ± 1.8 | 2.4 ± 1.3 |
| Total person-years | 859 | 2,245 | 3,988 | 4,209 | 504 | 291 |
| Psychological distress |  |  |  |  |  |  |
| CES-D 10 score (max:30; mean ± SD) | 5.5 ± 4.2 | 5.8 ± 4.8 | 4.9 ± 4.0 | 5.3 ± 4.3 | 5.9 ± 4.3 | 6.4 ± 4.3 |
| CES-D 10 ≥ 10, N (%) | 17 (14.5) | 53 (19.9) | 39 (9.4) | 54 (12.5) | 22 (18.0) | 22 (23.2) |
| CES-D 10 ≥ 11, N (%) | 14 (12.0) | 42 (15.7) | 32 (7.7) | 43 (10.0) | 12 (9.8) | 15 (15.8) |
| CES-D 10 ≥ 12, N (%) | 13 (11.1) | 34 (12.7) | 24 (5.8) | 33 (7.6) | 10 (8.2) | 9 (9.5) |
| Work expectation |  |  |  |  |  |  |
| Mean ± SD | 69.3 ± 24.8 | 76.1 ± 21.8 | 75.0 ± 24.1 | 78.1 ± 21.3 | 65.7 ± 24.1 | 71.0 ± 19.7 |
| Median (interquartile range) | 70 (40) | 80 (40) | 80 (40) | 80 (30) | 70 (30) | 80 (27.5) |
| Age (mean ± SD) | 49.8 ± 1.9 | 49.9 ± 2.0 | 49.9 ± 2.0 | 49.9 ± 2.0 | 50.0 ± 1.9 | 49.9 ± 2.2 |
| Female, N (%) | 85 (72.6) | 203 (75.5) | 137 (32.8) | 134 (31.0) | 95 (77.9) | 91 (94.8) |
| Rurality, N (%) | 19 (16.2) | 68 (25.3) | 43 (10.3) | 87 (20.1) | 22 (18.0) | 16 (16.7) |
| Education, N (%) |  |  |  |  |  |  |
| Lower secondary | 41 (35.0) | 124 (46.1) | 101 (24.2) | 100 (23.1) | 49 (40.2) | 33 (34.4) |
| Upper secondary, vocational training | 71 (60.7) | 118 (43.9) | 208 (49.8) | 261 (60.4) | 64 (52.5) | 58 (60.4) |
| Tertiary | 5 (4.3) | 27 (10.0) | 109 (26.1) | 71 (16.4) | 9 (7.4) | 5 (5.2) |
| Married/partnered, N (%) | 102 (87.2) | 246 (91.4) | 384 (91.9) | 398 (92.1) | 106 (86.9) | 86 (89.6) |
| Household income (in 10,000 Korean won), N(%) |  |  |  |  |  |  |
| < 1,620 | 47 (40.2) | 94 (34.9) | 48 (11.5) | 95 (22.0) | 45 (36.9) | 35 (36.5) |
| 1,620 - 2,879 | 29 (24.8) | 75 (27.9) | 94 (22.5) | 110 (25.5) | 30 (24.6) | 26 (27.1) |
| 2,880 - 4,235 | 28 (23.9) | 62 (23.0) | 130 (31.1) | 119 (27.5) | 27 (22.1) | 20 (20.8) |
| ≥ 4,236 | 13 (11.1) | 38 (14.1) | 146 (34.9) | 108 (25.0) | 20 (16.4) | 15 (15.6) |
| Total wealth (in 10,000 Korean won), N(%) |  |  |  |  |  |  |
| < 4,050 | 50 (42.7) | 77 (28.7) | 88 (21.1) | 87 (20.3) | 38 (31.1) | 24 (25.0) |
| 4,050 - 10,316 | 25 (21.4) | 71 (26.5) | 102 (24.5) | 107 (25.0) | 32 (26.2) | 26 (27.1) |
| 10,316 - 21,790 | 21 (17.9) | 59 (22.0) | 124 (29.7) | 112 (26.2) | 25 (20.5) | 19 (19.8) |
| ≥ 21,790 | 21 (17.9) | 61 (22.8) | 103 (24.7) | 122 (28.5) | 27 (22.1) | 27 (28.1) |
| Self-rated health |  |  |  |  |  |  |
| Poor/fair | 41 (35.0) | 83 (30.9) | 89 (21.3) | 99 (22.9) | 47 (38.5) | 24 (25.0) |
| Good | 58 (49.6) | 143 (53.2) | 239 (57.2) | 247 (57.2) | 59 (48.4) | 59 (61.5) |
| Very good/excellent | 18 (15.4) | 43 (16.0) | 90 (21.5) | 86 (19.9) | 16 (13.1) | 13 (13.5) |
| At least one chronic condition:<br>hypertension, diabetes, cancer, chronic<br>lung disease, heart problem, stroke,<br>arthritis, N(%) | 25 (21.4) | 56 (20.8) | 100 (23.9) | 102 (23.6) | 38 (31.1) | 23 (24.0) |
| Current smoker, N(%) | 24 (20.5) | 42 (15.6) | 124 (29.7) | 151 (35.0) | 19 (15.6) | 3 (3.1) |
| Current drinker, N(%) | 49 (41.9) | 103 (38.3) | 266 (63.6) | 277 (64.1) | 49 (40.2) | 30 (31.2) |

**Supplementary Table 6.** Adjusted odd ratios and 95% confidence intervals for the association between continuous work expectations and psychological distress by employment transition in the United States using alternative threshold scores for CES-D 8 (CES-D 8 ≥ 3 and ≥ 5)

|  | Odds ratio (95% confidence interval) for psychological distress |  |  |
| --- | --- | --- | --- |
|  | Pre-recession<br>(2006 - Nov 2007) | Recession<br>(Dec 2007 - Jun 2009) | Recovery<br>(July 2009 - 2018) |
| **CES-D 8 $\geq 3$** |  |  |  |
| Full sample (N = 2,647) | 1.01 (0.97, 1.05) | 0.94 (0.91, 0.98)* | 0.96 (0.93, 0.99)* |
| Entrance to full-time (N = 122) | 1.26 (1.05, 1.51)* | 0.89 (0.73, 1.09) | 0.99 (0.88, 1.12) |
| Precarious employment (N = 361) | 1.00 (0.90, 1.11) | 0.94 (0.84, 1.05) | 0.97 (0.90, 1.06) |
| Stable full-time (N = 884) | 1.04 (0.96, 1.12) | 0.98 (0.91, 1.07) | 0.97 (0.92, 1.02) |
| Self-employment, transition to self-employment (N = 275) | 1.01 (0.89, 1.14) | 1.12 (0.97, 1.29) | 0.98 (0.91, 1.06) |
| Exit labor force (N = 414) | 0.99 (0.91, 1.09) | 0.98 (0.89, 1.08) | 0.99 (0.91, 1.07) |
| Disabled, retired, out of labor force (N = 591) | 1.08 (0.96, 1.23) | 0.99 (0.83, 1.18) | 0.99 (0.88, 1.12) |
| **CES-D 8 $\geq 5$** |  |  |  |
| Full sample (N = 2,647) | 0.98 (0.94, 1.03) | 0.93 (0.88, 0.98)* | 0.94 (0.90, 0.98)* |
| Entrance to full-time (N = 122) | 1.09 (0.79, 1.50) | 0.77 (0.53, 1.12) | 0.93 (0.77, 1.11) |
| Precarious employment (N = 361) | 0.97 (0.86, 1.11) | 0.90 (0.79, 1.03) | 0.94 (0.85, 1.04) |
| Stable full-time (N = 884) | 0.93 (0.81, 1.06) | 1.08 (0.95, 1.25) | 0.94 (0.87, 1.03) |
| Self-employment, transition to self-employment (N = 275) | 1.15 (0.94, 1.41) | 1.24 (0.98, 1.57) | 1.08 (0.96, 1.22) |
| Exit labor force (N = 414) | 0.95 (0.85, 1.07) | 0.95 (0.83, 1.08) | 0.94 (0.83, 1.06) |
| Disabled, retired, out of labor force (N = 591) | 1.01 (0.87, 1.18) | 0.89 (0.70, 1.11) | 0.85 (0.71, 1.01) |

<sup>a</sup> All models adjusted for centered age, sex, minority status, education, marital status, household income, total wealth, self-rated health, having at least one chronic condition, random intercept, and random slope for age
<sup>b</sup> 95% confidence intervals were approximated using the delta method
\* $p < 0.05$

**Supplementary Table 7.**
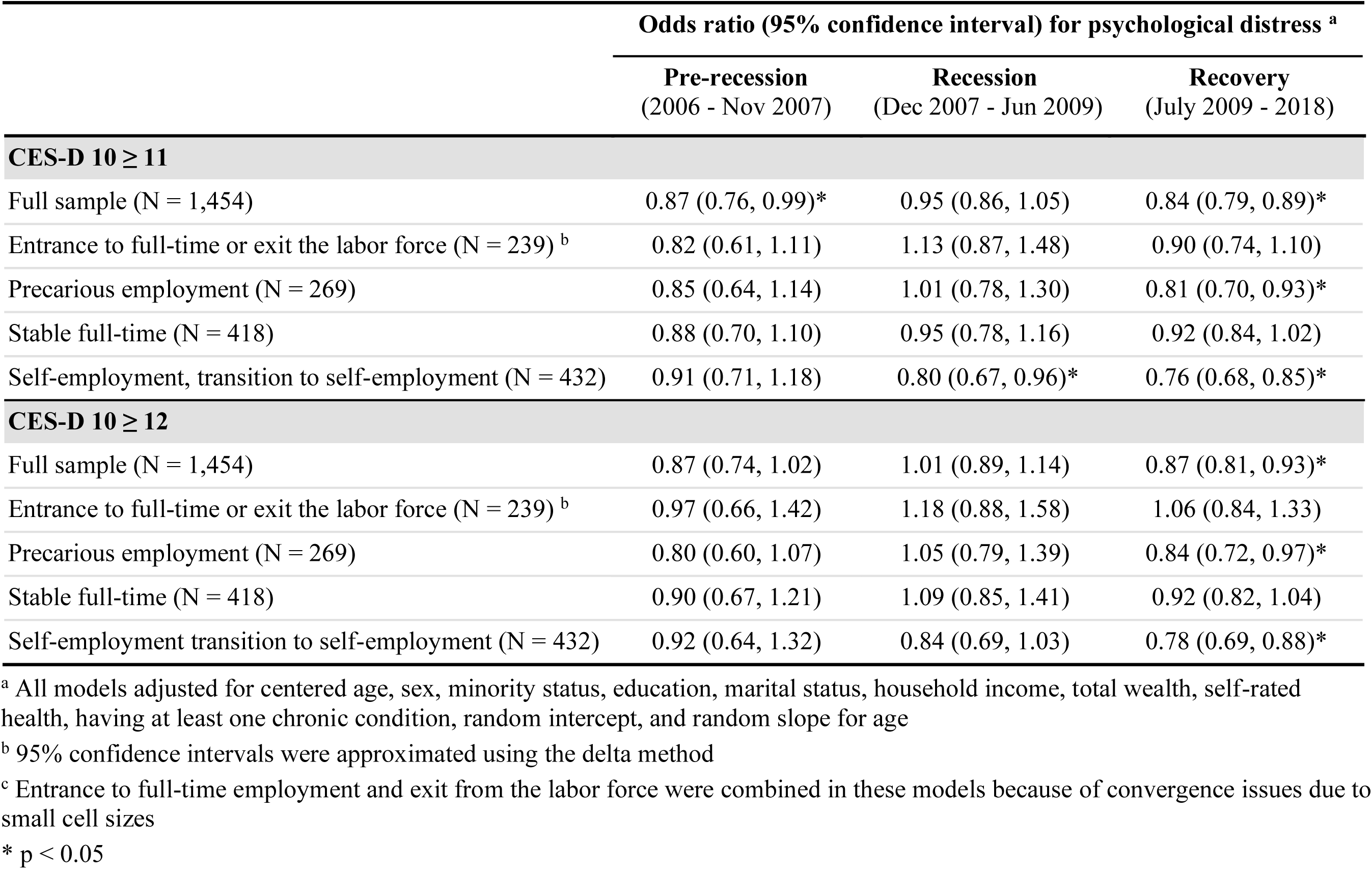
Adjusted odd ratios and 95% confidence intervals for the association between continuous work expectations and psychological distress by employment transition in South Korea using alternative threshold scores for CES-D 10 (CES-D 10 ≥ 11 and ≥ 12)

**Supplementary Table 8.** Adjusted odd ratios and 95% confidence intervals for the association between work expectations and psychological distress by employment transition among currently working respondents in the United States

| | Odds ratio (95% confidence interval) for CESD-8 $\geq 4$ <sup>a</sup> | | |
| --- | --- | --- | --- |
|  | Pre-recession<br>(2006 - Nov 2007) | Recession<br>(Dec 2007 - Jun 2009) | Recovery<br>(July 2009 - 2018) |
| **United States** |  |  |  |
| Full sample (N = 2,100) | 1.01 (0.94, 1.08) | 1.00 (0.93, 1.08) | 0.97 (0.92, 1.02) |
| Entrance to full-time (N = 121) | 2.16 (1.18, 3.95)* | 0.65 (0.42, 1.01) | 1.10 (0.86, 1.40) |
| Precarious employment (N = 348) | 1.04 (0.78, 1.37) | 1.05 (0.81, 1.35) | 0.74 (0.57, 0.96)* |
| Stable full-time (N = 883) | 0.97 (0.85, 1.10) | 0.99 (0.87, 1.13) | 0.97 (0.89, 1.05) |
| Self-employment, transition to self-employment (N = 275) | 1.01 (0.87, 1.18) | 1.22 (1.01, 1.48)* | 0.99 (0.88, 1.12) |
| Exit labor force (N = 398) | 0.98 (0.88, 1.09) | 0.94 (0.80, 1.10) | 1.01 (0.87, 1.17) |

<sup>a</sup> Work expectation was modeled as a continuous variable and scaled to 0-10. All models adjusted for centered age, sex, minority status, education, marital status, household income, total wealth, self-rated health, having at least one chronic condition, random intercept, and random slope for age
<sup>b</sup> 95% confidence intervals were approximated using the delta method
<sup>c</sup> Model did not converge among respondents who were retired or not in the labor force
\* $p < 0.05$

**Supplementary Table 9.** Adjusted odd ratios and 95% confidence intervals for the association between categorical work expectations and psychological distress by employment transition in the United States

|  | **Odds ratio (95% confidence interval) for CESD-8 $\geq 4$ <sup>a</sup>** |  |  |  |  |  |
| --- | --- | --- | --- | --- | --- | --- |
|  | **Full sample** | **Entrance to full-time** | **Precarious employment** | **Stable full-time** | **Self-employment transition to self-employment** | **Exit labor force** |
|  | (n = 2,647) | (n = 122) | (n = 361) | (n = 884) | (n = 275) | (n = 414) |
| *<u>Pre-recession (2006 - November 2007)</u>* |  |  |  |  |  |  |
| 0-40% | 0.91<br>(0.57, 1.45) | 1.45<br>(0.11, 19.71) | 0.49<br>(0.14, 1.69) | 0.56<br>(0.19, 1.62) | 0.48<br>(0.08, 2.97) | 0.84<br>(0.28, 2.51) |
| 41-60% | ref | ref | ref | ref | ref | ref |
| 61-89% | 0.68<br>(0.35, 1.31) | 4.22<br>(0.11, 155.9) | 0.10<br>(0.02, 0.58)* | 0.60<br>(0.18, 2.02) | 4.50<br>(0.56, 36.16) | 0.86<br>(0.19, 3.98) |
| 90-100% | 1.04<br>(0.59, 1.85) | 15.24<br>(0.80, 289.7) | 0.80<br>(0.19, 3.31) | 0.61<br>(0.20, 1.83) | 0.93<br>(0.14, 6.29) | 0.64<br>(0.16, 2.56) |
| *<u>Recession (December 2007 - June 2009)</u>* |  |  |  |  |  |  |
| 0-40% | 2.25<br>(1.34, 3.78)* | 2.13<br>(0.22, 20.54) | 1.64<br>(0.51, 4.25) | 0.82<br>(0.25, 2.66) | 2.20<br>(0.26, 18.38) | 4.25<br>(1.21, 14.94)* |
| 41-60% | ref | ref | ref | ref | ref | ref |
| 61-89% | 1.52<br>(0.77, 3.00) | 0.17<br>(0.01, 3.96) | 0.61<br>(0.12, 3.22) | 1.57<br>(0.46, 5.32) | 9.41<br>(0.99, 89.8) | 4.68<br>(0.91, 24.07) |
| 90-100% | 1.36<br>(0.70, 2.64) | 0.15<br>(0.003, 6.72) | 1.05<br>(0.25, 4.52) | 0.76<br>(0.22, 2.62) | 18.25<br>(1.98, 168.5)* | 1.11<br>(0.16, 7.72) |
| *<u>Recovery (July 2009 - 2018)</u>* |  |  |  |  |  |  |
| 0-40% | 1.75<br>(1.22, 2.50)* | 3.22<br>(0.76, 13.67) | 2.12<br>(0.90, 4.98) | 1.34<br>(0.69, 2.59) | 0.83<br>(0.30, 2.25) | 3.54<br>(1.03, 12.18)* |
| 41-60% | ref | ref | ref | ref | ref | ref |
| 61-89% | 1.15<br>(0.70, 1.90) | 2.82<br>(0.43, 18.39) | 1.01<br>(0.29, 3.45) | 1.18<br>(0.50, 2.75) | 0.73<br>(0.20, 2.64) | 5.18<br>(0.96, 27.95) |
| 90-100% | 1.72<br>(1.04, 2.85)* | 6.23<br>(0.82, 47.34) | 2.54<br>(0.74, 8.73) | 1.15<br>(0.47, 2.81) | 2.36<br>(0.68, 8.26) | 2.41<br>(0.40, 14.49) |

<sup>a</sup> All models adjusted for centered age, sex, minority status, education, marital status, household income, total wealth, self-rated health, having at least one chronic condition, random intercept, and random slope for age
<sup>b</sup> 95% confidence intervals were approximated using the delta method
<sup>c</sup> Model did not converge among respondents who were retired or not in the labor force
\* $p < 0.05$

**Supplementary Table 10.**
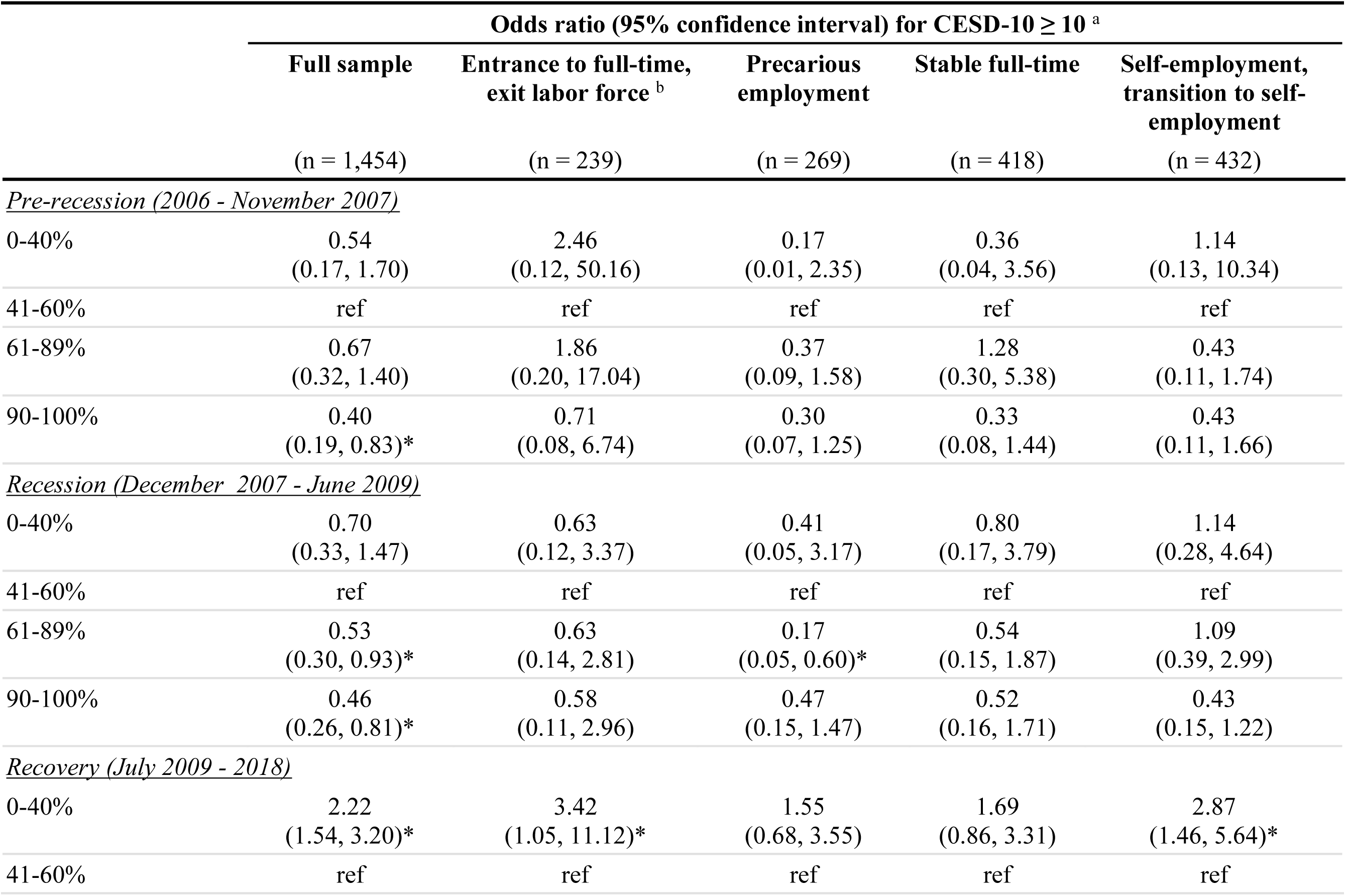
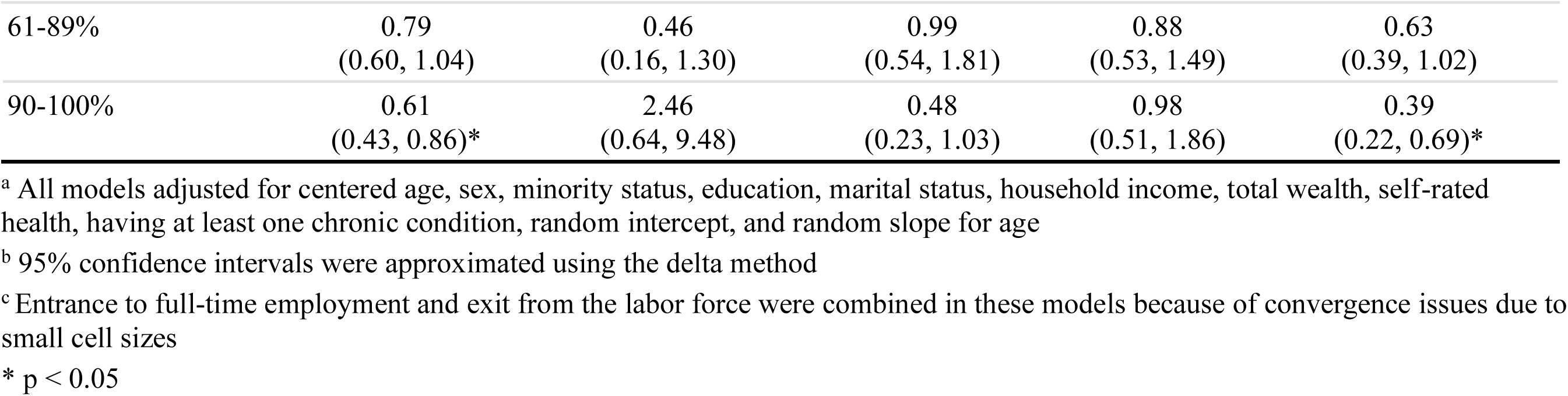
Adjusted odd ratios and 95% confidence intervals for the association between categorical work expectations and psychological distress by employment transition in South Korea

